# Dementia risk factors in former contact sports participants: prospective cohort study

**DOI:** 10.1101/2024.01.15.24301327

**Authors:** G. David Batty, Steven Bell, Urho M. Kujala, Seppo J. Sarna, Jaakko Kaprio

**Author notes:** Corresponding author: David Batty, Department of Epidemiology and Public Health, University College London, 1-19 Torrington Place, London, UK, WC1E 6BT. E. Role of the funding source: This report had no direct funding. All authors had final responsibility for the decision to submit for publication. Contributions: GDB generated the idea for the paper; formulated the plan for data analyses; prepared the tables; and drafted the manuscript. SB prepared figures and edited the manuscript. UMK and SJS initiated the cohort study and designed data collection; accessed and verified the data; and edited the manuscript. JK designed data collection; formulated the plan for data analyses; accessed, verified, and analysed the data; and edited the manuscript. Data sharing statement: *Bona fide* interested parties should contact UMK and SJS regarding data access. Declaration of Interest: None.

## Abstract

**Background:** The elevated dementia incidence in retired contact sport participants might be explained by a higher prevalence of established risk factors for the disease relative to the general population.

**Methods:** In this cohort study, former elite participants active between 1920 and 1965 in soccer (N=303), boxing (N=281), and wrestling (N=318) were recruited using sports yearbooks and records of sports associations. Men in a population control group were identified using records from a compulsory medical examination (N=1712). All study members were linked to hospital registers (1970-2015) and self-completion questionnaires were circulated (1985, 1995) from which we captured data on nine established risk factors for dementia: hypertension and diabetes status, alcohol intake, loneliness, depressive symptoms, cigarette smoking, body weight, educational attainment, and physical activity.

**Results:** There was little suggestion that former participants in contact sports had a higher prevalence of dementia risk factors relative to the general population. Rather, the balance of evidence was for more favourable risk factor levels in former athletes, as was particularly evident for ever having smoked cigarettes (range in odds ratios [95% confidence interval]: 0.32 [0.21, 0.48] for wrestling to 0.52 [0.36, 0.75] for soccer) and leisure-time physical activity (range in beta coefficients [95% confidence interval]: 1.34 [0.66, 2.02] for soccer to 1.80 [1.07, 2.52] for boxing).

**Conclusions:** The increased dementia rates in retired contact sport participants evident in epidemiological studies is unlikely to be explained by the risk factors examined here. This implicates other characteristics of contact sports, including a history of repeated head impact.

## Introduction

Recent epidemiological evidence suggests that retired contact sports participants have elevated rates of dementia relative to the general population.^1^ That these activities are characterised by head contact with the opponent (e.g., boxing, wrestling) and/or equipment (e.g., ‘heading’ the ball in soccer) raises the possibility that accumulated sub-concussive and concussive brain injury has a role in dementia occurrence.^2^

The cohort studies on which these findings are based have been generated from linked administrative records. While this has led to some large, well-powered datasets,^3,4^ they typically lack collateral data on established risk factors for dementia which are potential confounding factors in this context. If these risk factors, which include heavy alcohol intake, cigarette smoking, diabetes, and hypertension^5^ – some of which are also important health outcomes in their own right – are shown to be more prevalent in former contact sports participants, it raises the possibility that they are, in fact, responsible for raising dementia risk and not head impact itself.

Retired American footballers appear to have a higher prevalence of obesity and elevated blood pressure relative to the general population^6^ but whether this observation is generalisable to other contact sports is uncertain. Indeed, erstwhile elite soccer players have a lower incidence of both alcohol-related disorders^7^ and depression^8^ at follow-up. Given the clear paucity of data on dementia risk factors in former contact sports participants, using general population controls as a comparator, we examined levels of nine dementia risk factors amongst retired elite boxers, wrestlers, and soccer players – all sports which have previously been linked to a higher burden of dementia.^1,3,4^

## Methods

The present cohort study was initiated in 1978 to examine the life expectancy of former elite level athletes.^9-11^ In brief, this cohort comprises male retired athletes who represented Finland between 1920 and 1965 at least once in intercountry competitions (e.g., Olympic games, World or European championships). Full name, and place and date of birth were retrospectively extracted from sports yearbooks and registers of sports associations. Of the range of retired sports participants identified during this process, we included soccer players (N=303, mean age in 1978: 57.1 years; boxers (N=281, 56.4 years); and wrestlers (N=318, 60.9 years). A general population-based comparison group of 1712 men (55.2 years) who were in the same age cohort and area of residence as the athletes was retrospectively selected using a population-wide conscription dataset. When aged 20, these controls were classified as healthy at the compulsory medical examination for induction into military/civic service. All data collection was approved by the ethics committee of the Hospital Districts of Helsinki and Uusimaa, and all participants consented. The composition of this manuscript conforms to the Strengthening the Reporting of Observational Studies in Epidemiology (STROBE) Statement.^12^

### Assessment of dementia risk factors at follow-up

All study members were followed for the occurrence of hypertension and diabetes between 1970, when hospital records began in Finland (mean age athletes 45.4 years; controls 44.3 years), and 2015. In 1985, a questionnaire was mailed to surviving contact sports athletes (434 [83.5%] responded) and population controls (777 [76.9%] responded).

In the 1985 questionnaire, standard enquiries were made regarding cigarette smoking (ever smoked versus never); educational attainment (years); marital status (single, divorced, widowed vs married, remarried, cohabiting); and height and weight. Depressive symptoms were ascertained using the Brief Symptom Inventory mailed in 1995. Episodic heavy drinking was defined as more than five standard drinks (12g ethanol/drink) on a single occasion at least once a month, while passing out was denoted by alcohol-induced loss of consciousness at least once in the preceding year.^13^ A leisure-time physical activity index, expressed as metabolic equivalents (MET) per day, was the product of intensity, duration, and frequency of an array of activities.^14^ Feelings of loneliness were rated on a 3 point scale (not feeling lonely vs fairly lonely, very lonely). Questionnaire data in combination with information from the Central Population Registry were used to generate a variable for longest held occupation, our indicator of socioeconomic status.

Collection of these data provided nine of the twelve known dementia risk factors,^5^ some with multiple indicators: hypertension, diabetes, heavy alcohol intake (amount, frequency, passing out), social isolation (single marital status, loneliness), depression symptoms, cigarette smoking, higher body mass index, low educational attainment, and physical inactivity.

### Statistical analyses

Logistic (binary outcomes) and linear (continuous outcomes) regression were used to summarise the relationship between contact sports participation and dementia risk factors. In preliminary analyses, effect estimates adjusted for age were very similar to those additionally adjusted for socioeconomic status, so the results from the multiply-adjusted analyses are presented here. The only exception was for education which had an unsurprisingly high degree of collinearity with socioeconomic status (r=0.63); coefficients were therefore age-adjusted only. Analyses were computed using Stata 15 (StataCorp, College Station, TX) between October and December 2023.

## Results

In Figure 1 (categorical outcome variables) and Figure 2 (continuous) we show levels of dementia risk factors at follow-up in former participants in the three contact sports relative to general population controls. Collectively, there was very little suggestion of an increased level of these risk factors in retired athletes. Indeed, the general pattern of association was null, conclusions that were unchanged after applying a Bonferroni correction for multiple comparisons. Where statistical significance did occur, this was most frequently evident for more favourable risk factor levels in former athletes.

**Figure 1.**
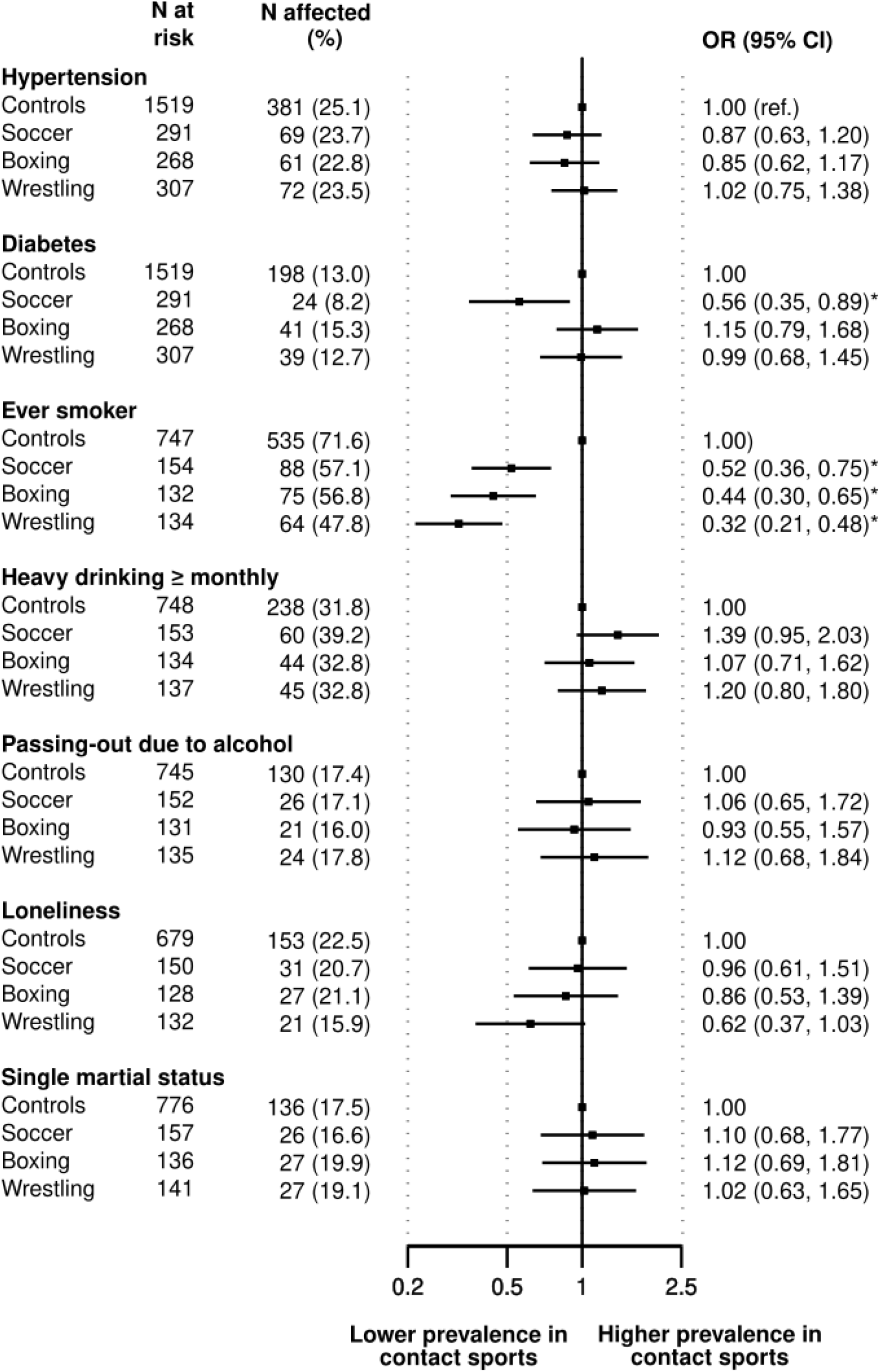
Odds ratios (95% confidence interval) for the prospective association between prior participation in contact sports (1920-1965) and dementia risk factors (1970-2015) Odds ratios are adjusted for age and socioeconomic status. Asterisk denotes statistical significance at p<0.05.

**Figure 2.**
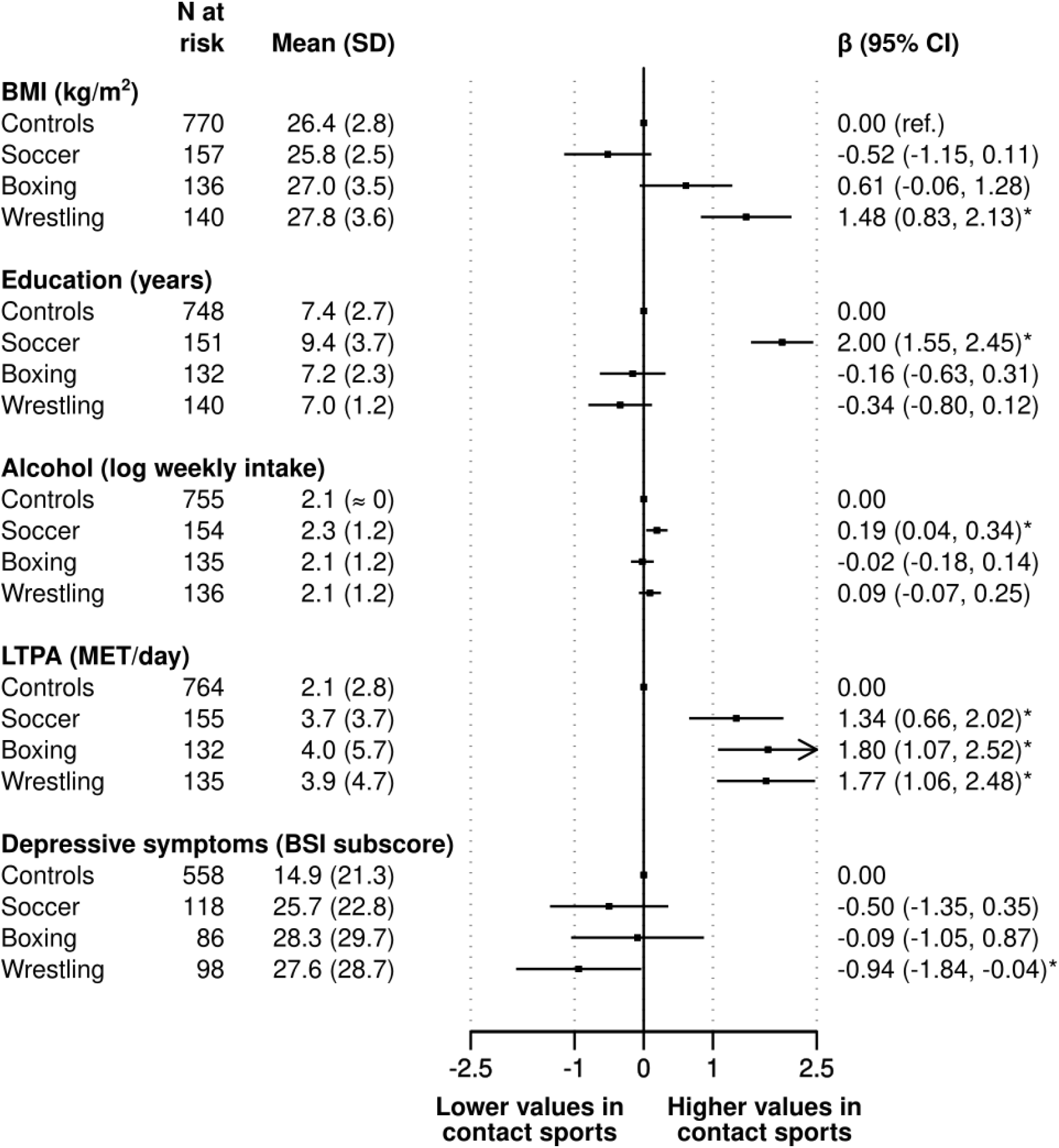
Beta coefficients (95% confidence interval) for the prospective association between prior participation in contact sports (1920-1965) and dementia risk factors (1970-2015) BMI, body mass index; LTPA, leisure-time physical activity; MET, metabolic equivalent; BSI, Brief Symptom Inventory. Beta coefficients are adjusted for age and socioeconomic status, except for education which is age-adjusted only. Asterisk denotes statistical significance at p<0.05. Previously published,^8^ the results for depression are shown here for the purposes of comparison with other dementia risk factors.

In comparison to population controls, we found a lower prevalence of ever having smoked cigarettes in all contact sports groups (range in odds ratios [95% confidence interval]: 0.32 [0.21, 0.48] for wrestling to 0.52 [0.36, 0.75] for soccer), alongside a lower risk of diabetes in retired soccer players only (0.56 [0.35, 0.89]). All former contact sports participants also had higher levels of post-career physical activity (range in beta coefficients [95% confidence interval]: 1.34 [0.66, 2.02] for soccer to 1.80 [1.07, 2.52] for boxing), while retired soccer players were better educated (1.21 [0.84, 1.58]) and past participants in wrestling reported fewer depressive symptoms than controls (−0.94 [-1.84, -0.04]). The only instances of elevated levels of dementia risk factors in former contact athletes were for alcohol intake in soccer players (0.19 [0.04, 0.34]) and higher body mass index in wrestlers (1.48 [0.82, 2.14]).

## Discussion

The main finding of this cohort study was that there was very little evidence that former participants in elite-level contact sports had a higher prevalence of dementia risk factors at follow-up relative to the general population. Indeed, nine of eleven statistically significant associations were for protective risk factor levels in former athletes and this was particularly evident for the health behaviours of cigarette smoking and physical activity.

The present findings suggest that dementia risk factors captured in the present study do not explain the elevated rates of the condition in cohort studies of contact sport participants. Rather, it is more likely that the poorer brain health is ascribed to other characteristics which may include recurrent head impacts, a suggestion that has some biological plausibility.^15^ Given the paucity of studies of dementia risk factors in former athletes, particularly for the indices not measured herein – hearing impairment and air pollution^5^ – our findings need to be tested in different contexts, particularly in women as their increasing participation in contact sports such as soccer has potential consequences for the future burden of neurodegenerative disease.

## Data Availability

Bona fide interested parties should contact UMK and SJS regarding data access.

## Acknowledgements

The preparation of this manuscript was unfunded. GDB is supported by the UK Medical Research Council (MR/P023444/1; MR/X003434/1) and the US National Institute on Aging (1R56AG052519-01; 1R01AG052519-01A1); SB by Cancer Research UK (A27657); and JK by the Academy of Finland Centre of Excellence in Complex Disease Genetics (336823 & 352792).

